# Healthy Family Program on population blood pressure: a multicenter, parallel group, cluster randomized, controlled trial in rural China (Healthy Family Program)

**DOI:** 10.1101/2025.04.07.25325383

**Authors:** Xiaolei Lin, Yangyang Tang, Chao Jiang, Jun Cai, Craig S. Anderson, Xin Du

**Affiliations:** School of Data Science, Fudan University, Shanghai, China; Beijing Anzhen Hospital, Capital Medical University, Beijing, China; The George Institute for Global Health, University of New South Wales, Sydney, Australia; The Institute of Science and Technology for Brain-inspired Intelligence, Fudan University, Shanghai, China; Heart Health Research Center, Beijing, China

**Keywords:** Blood pressure management, Cluster randomized trial, Multifaceted interventions, Rural China, Family health instructor, Statistical analysis plan

## Abstract

The Healthy Family Program trial aims to evaluate the effectiveness of a community-based, health instructor led, multifaceted family intervention, as compared with usual care, on blood pressure (BP) management among Chinese rural residents, with or without hypertension. It is designed as a two-arm, parallel, cluster randomized trial of 80 villages for 12 months. This statistical analysis plan pre-specifies the method of analysis for every outcome and key variables conducted in the trial.

The primary outcome is change in systolic BP from baseline to 6 months in all participants, reported as the absolute difference between intervention and control groups. The primary outcome will be modelled using a linear mixed effect model based on a participant-level analysis. The model will include random effects at the village and family level to account for the clustering of participants within village and within family, respectively.

**Revision Summary:** This version contains this updated information.

1. We modified the primary analysis to account for potential correlation of outcomes within families. A mixed effect model was extended with the same set of fixed effects, village random effect, and family random effect (nested within village). However, since the number of participants within a family is relatively small, the estimation of family random effect could be numerically unstable. When the variance of the family random effect could not be estimated, a mixed effect model with village random effect will be conducted instead.
2. Additional sensitivity analysis is planned at the family level, where family-level mean change in systolic blood pressure will be analyzed using a linear mixed effect model.
3. Additional subgroup analyses is included by stratifying participants based on family size and township socio-economic level.
4. In the subgroup analysis, the categorization of the Mini-EAT dietary scale was modified from the original cutoffs of <60 and ≥60 points to new thresholds based on the sample medium (i.e., below the median vs. equal to or above the median).

## 2 Administrative information

### 2.1 Study identifiers

- ClinicalTrial.gov register Identifier: NCT06427096, Date: 18 May 2024

#### 2.2 Revision history

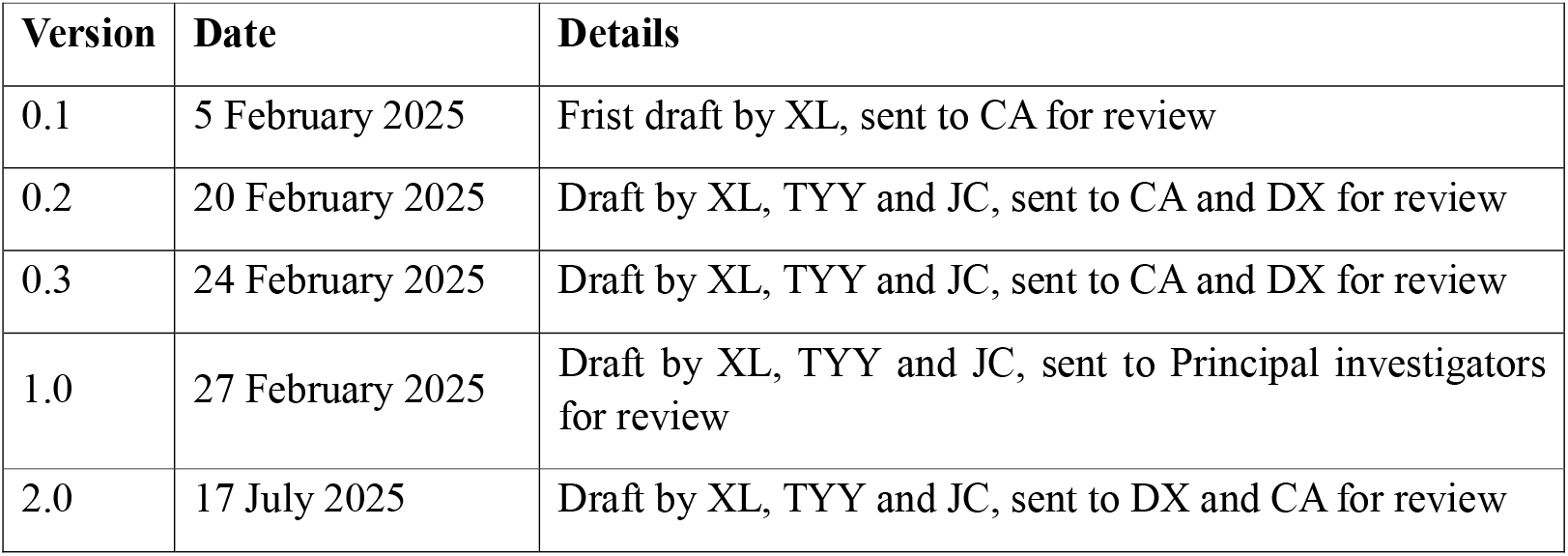

| Version | Date | Details |
| --- | --- | --- |
| 0.1 | 5 February 2025 | Frist draft by XL, sent to CA for review |
| 0.2 | 20 February 2025 | Draft by XL, TYY and JC, sent to CA and DX for review |
| 0.3 | 24 February 2025 | Draft by XL, TYY and JC, sent to CA and DX for review |
| 1.0 | 27 February 2025 | Draft by XL, TYY and JC, sent to Principal investigators for review |
| 2.0 | 17 July 2025 | Draft by XL, TYY and JC, sent to DX and CA for review |

#### 2.3 Contributors to the statistical analysis plan

##### 2.3.1 Roles and responsibilities

2.3 Contributors to the statistical analysis plan 2.3.1 Roles and responsibilities
| Name and ORCID | Affiliation | Role on study | SAP contribution |
| --- | --- | --- | --- |
| Prof. Xiaolei Lin | Fudan University | Statistician | Prepare initial draft and subsequent versions |
| Dr. Yangyang Tang | Beijing Anzhen Hospital, Capital Medical University | Study coordinator | Reviewed every draft and approved final version |
| Prof. Chao Jiang | Beijing Anzhen Hospital, Capital Medical University | Principal investigator | Reviewed every draft and approved final version |
| Prof. Xin Du | Beijing Anzhen Hospital, Capital Medical University | Principal investigator | Reviewed every draft and approved final version |
| Prof. Craig Anderson | Fudan University | Principal investigator | Reviewed every draft and approved final version |
| Prof. Jun Cai | Beijing Anzhen Hospital, Capital Medical University | Principal investigator | Reviewed every draft and approved final version |

##### 2.3.2 Approvals

The undersigned have reviewed this plan and approve it as final. They find it to be consistent with the requirements of the protocol as it applies to their respective areas. They also find it to be compliant with International Conference on Harmonisation (ICH-E9) principles and in particular, confirm that this analysis plan was developed in a completely blinded manner (i.e. without knowledge of the effect of the intervention[s] being assessed)

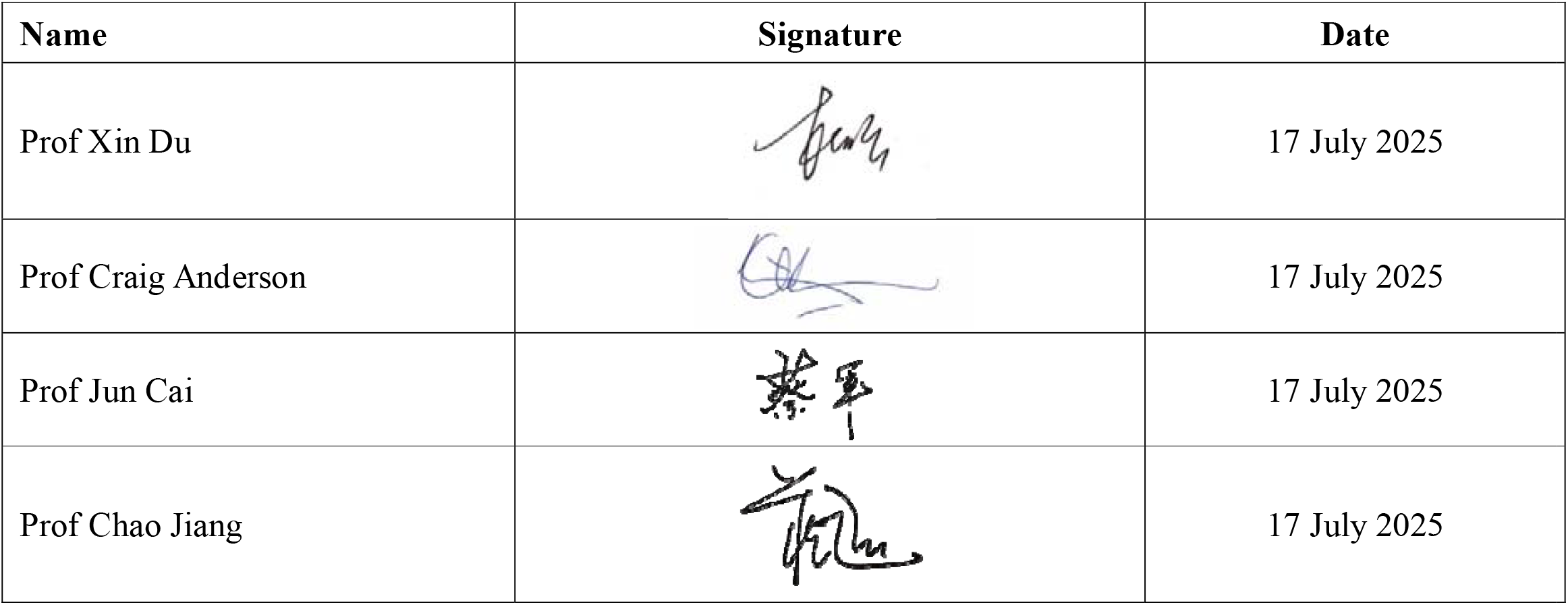

| Name | Signature | Date |
| --- | --- | --- |
| Prof Xin Du |  | 17 July 2025 |
| Prof Craig Anderson |  | 17 July 2025 |
| Prof Jun Cai |  | 17 July 2025 |
| Prof Chao Jiang |  | 17 July 2025 |

## 3 Introduction

### 3.1 Study synopsis

The Healthy Family Program is an investigator-initiated and conducted, multicenter, open-label, parallel-group, blinded outcome assessed, cluster randomized controlled trial being undertaken in 80 villages (each with approximately 100 residents) with a target to enroll a total of 8000 older adults (aged 40-80 years). The aim is to determine whether compared to a usual care control group, a multifaceted intervention led by family health instructors can reduce BP in rural village residents, regardless of their hypertension status, over a 6-month intervention and 12-month follow-up period. In this trial, the unit of the cluster is the village. Villages were randomly assigned (1:1) to the intervention or control group using a centralized computerized system, with randomization occurring after screening for eligible households and residents, and stratified by township (7 townships total). The enrollment period was from May 23, 2024 to July 18, 2024. Villages allocated to the intervention group received a family health instructor intervention focused on six areas: education for a healthy lifestyle, free provision of sodium substitute, weight management, physical exercise, BP monitoring, and appropriate antihypertensive treatment for individuals with hypertension. Villages allocated to the control group received usual standard of care.

### 3.2 Study population

Ninety-six candidate villages were pre-screened based on the following criteria: (1) a minimum of 200 permanent households, and (2) at least 60 households with two or more members aged 40 years or more. These villages are located in the Ruyang County, Henan Province. The leaders of these villages were convened to introduce the research protocol. The final 80 villages selected to participate in the study were chosen based on their willingness and ability to participate. The flow diagram of recruitment and follow-up is shown in Figure 1.

**Figure 1.**
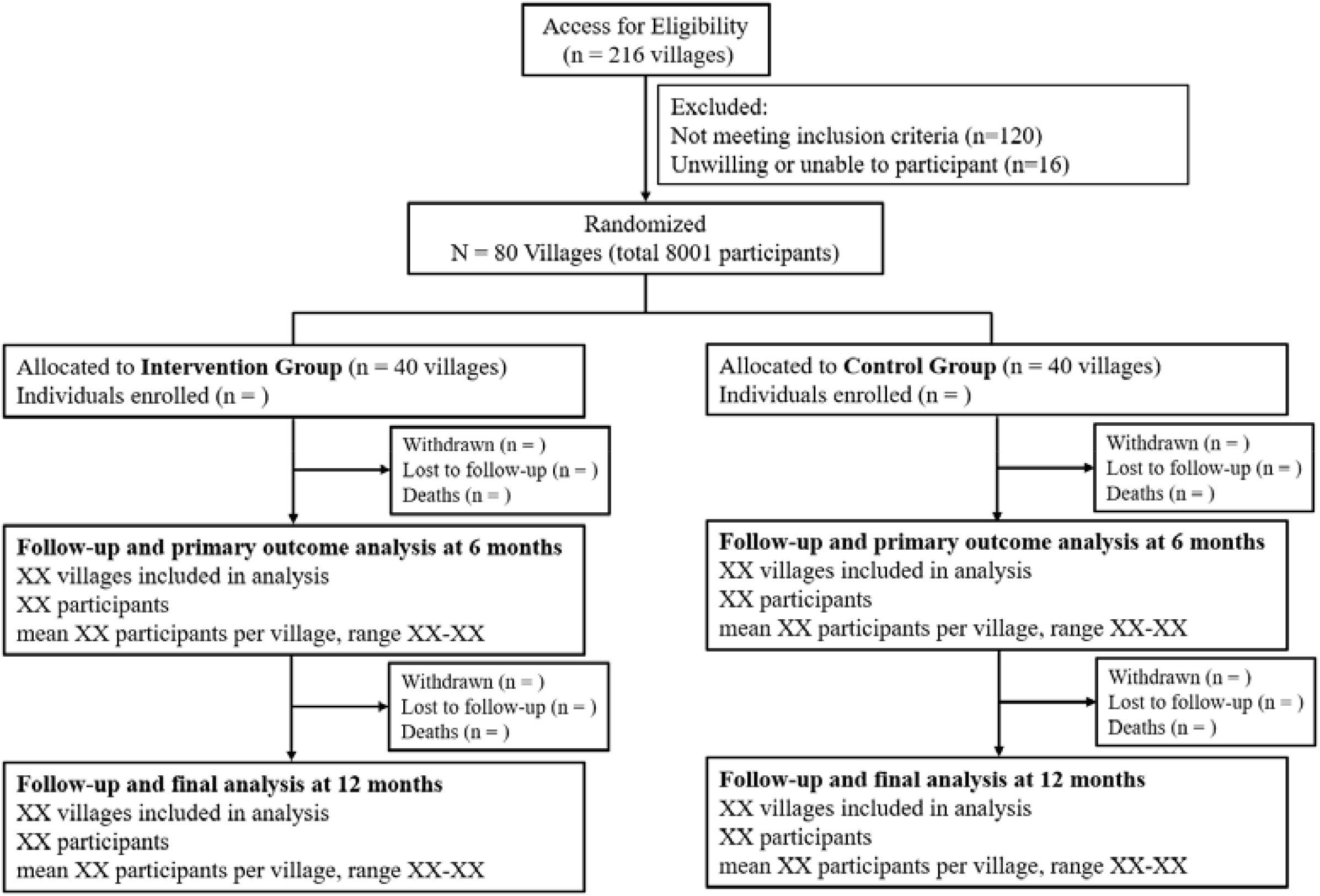
The flow diagram of Healthy Family Program.

For each participating village, the family health instructor was responsible for identifying 100 individuals from 30 to 50 households. Each household must have had at least two family members who agreed to participate in the study. Individuals were invited to participate in the study regardless of their BP level or history of antihypertensive treatment.

#### 3.2.1 Inclusion criteria

##### Village level

Village sites are eligible if they meet all following requirement:

- With a minimum of 200 permanent households
- At least 60 households with two or more members aged 40 years or more

##### Household level

Households are eligible if they meet all following requirement:

- At least 2 family members aged 40-80 years meet the inclusion and exclusion criteria of family members and willing to participate in this study
- At least one family member can use the smartphone to log their measurements (e.g. blood pressure and weight)

##### Individual level

Participants who lived in villages are eligible if they meet all the following requirement

- Aged 40-80 years old, regardless of their BP levels and antihypertensive treatment
- No travel plan for more than 1 month during the study period
- Able to written or fingerprinted informed consent form

#### 3.2.2 Exclusion criteria

##### Village level

Villages are not eligible if they meet any of the following criteria:

- Unwilling or unable to participant
- Participating in other clinical trials

##### Household level

Households are not eligible if they meet any of the following criteria:

- Any of the family member currently participating in any other hypertension-related trials
- The family health instructors determined the family is not suitable to participate

##### Individual level

Participants are not eligible if they meet any of the following criteria

- Significant cognitive dysfunction
- Advanced tumor, dialysis, or other serious diseases
- Bed-bound or unable to perform daily self-care tasks
- Diagnosed of secondary hypertension
- Having birth plans in the next six months, pregnant or lactating women
- Other ineligible circumstances judged by the investigators

## 4 Study interventions

### 4.1 Multifaceted intervention group

The main components of the multifaceted intervention strategies include: 1) Establishing a BP management team led by family health instructors, with team members including family leaders and village doctors, and 2) Implementing an intervention that focused on six areas: education for a healthy lifestyle, free provision of sodium substitute, weight management, physical exercise, BP monitoring, and appropriate antihypertensive treatment for individuals with hypertension. Participants in the intervention group will receive low-sodium salt, BP monitors, and weighing scales free of charge, provided at the household level. The BP management team worked collaboratively to promote a healthy lifestyle and better BP management in participants. Furthermore, several monitoring strategies, like documentation of activities, salt consumption monitoring, health data monitoring and site visits have been implemented to ensure intervention fidelity.

### 4.2 Control group

The control group will receive routine BP management without any additional educational interventions by a family health instructor or villager personnel. They will not be provided with low-sodium salt, BP monitors, and electronic weighing scales. However, they will be required to cooperate for data collection in the same manner as the intervention group regarding antihypertensive medication usage, lifestyle factors, and reporting of CVD events.

## 5 Outcomes

### 5.1 Primary outcome

The primary outcome is the change in SBP from baseline to 6 months in all participants for comparison between the intervention group and control group. BP is measured in all participants using the Microlife BP B2 Basic monitor, with a cuff specifically designed for this model (suitable for arm circumferences ranging from 22 to 42 cm). After a 5-minute rest, three measurements will be taken in one setting with a 1-minute interval, and the mean of the second and third BP readings will be used for all analyses, while the first is discarded.

### 5.2 Secondary outcomes

Secondary outcomes include:

- The change in SBP between the intervention and control groups at 12 months;
- The change in DBP between the intervention and control groups at 6 months;
- The proportion of participants with SBP≥130 or DBP≥80 mmHg at 6 month;
- The proportion of participants with antihypertensive therapy at 6 months;

### 5.3 Safety outcomes

All serious adverse events (SAEs) according to standard definitions and Adverse Events of Special Interest (AESIs) will be collected during follow up. AESIs include hypotension, syncope, hyponatremia, hyperkalemia, injurious falls, and acute kidney injury. The adverse events will be self-reported by participants, with clinical records used for confirmation.

## 6 Analysis principles

### 6.1 Sample size

Based on a trial conducted in China[1], and assuming an standard deviation of 19 mmHg for SBP and an intraclass correlation coefficient is 0.05, a sample size of 8000 individuals (4000 in the intervention group and 4000 in the control group) is estimated to provide 80% power (with a two-sided alpha of 0.05) to detect a between-group difference of ≥3 mmHg in mean SBP. For each village, one family health instructor and 100 family members are required to ensure consistent participant numbers across all villages. A total of 80 villages in Ruyang county will provide 8000 participants.

### 6.2 Software

Analyses will be conducted primarily using SAS Enterprise Guide (version 8.3 or above) and R (version 4.0.0 or above).

### 6.3 Interim analysis

No formal interim analyses were conducted during the study.

### 6.4 Multiplicity adjustment

Statistical tests are two-sided with a nominal level of 5%. Analyses of the primary outcome will be unadjusted for multiplicity. For all secondary outcomes, we will control the family-wise error rate by applying a sequential Holm-Sidak correction. Briefly, the approach consists of ordering all p-values from smallest to largest, and then comparing them to an adjusted level of significance calculated as 1-(1-0.05)^1/C^, where C indicates the number of comparisons that remain. The sequential testing procedure stops as soon as a p value fails to reach the corrected significance level. This will apply only to the primary analysis of an outcome (i.e. not to sensitivity analyses). No multiplicity adjustment will be applied to safety outcomes.

### 6.5 Timing of analysis

The primary outcome will be analyzed at 6 months. After the 12-month follow-up, we will conduct the final analysis.

### 6.6 Data sets analysed

All visits and data collection will be conducted by the trained CRCs, accompanied by family health instructors. Data collection occurs at baseline (post-randomization), 6 months (at the end of the intervention) and 12 months (at the end of follow-up) for all participating family members on a household basis. Data will be upload into a purpose-built electronic data capture (EDC) system.

#### 6.6.1 Analysis populations

The intention-to-treat (ITT) population: all randomized participants, irrespective of their diagnosis or adherence to the intervention protocol, were included, except for those with missing primary outcome data or who withdrew consent for data usage.

#### 6.6.2 Analyses strategy

All analyses and reporting of results will follow CONSORT guidelines for cluster randomized controlled trials[2]. The analysis of all primary and secondary outcomes will use a (generalized) linear mixed-effects model with village and family as the random effects, accounting for the clustering effect[3, 4]. The ITT analyses will be performed, where study outcomes will be compared across participants based on their village randomization, regardless of adherence to the intervention. The results for the primary outcome will be presented as mean difference with two-sided 95% CI. In case of excessive skewness and/or kurtosis, analytical methods for asymmetric data or transformation will be considered for continuous outcomes. A full likelihood model will be used to handle missing data. This model utilizes both observed and missing data, leading to improved efficiency and accuracy in estimating model parameters[5]. A two-sided P value <0.05 for the primary outcome will be considered statistically significant. All confidence interval (CI) will be at the 95% level.

The consistency of treatment effects on the primary outcome will be explored in subgroups. Interaction effects within the subgroups will be assessed by adding an interaction term into the model. The subgroup analyses will not be adjusted for multiple testing and will be considered exploratory.

## 7 Planned analyses

### 7.1 Subject disposition

The flow diagram of villages and participants through the trial will be presented in Figure 1, following the Consolidated Standards of Reporting Trials (CONSORT) for cluster-level studies[2]. The report will include the following: the number of centers assessed for eligibility, reasons for exclusion, the number of enrolled participants, and the number of participants who completed the baseline and follow-up visits in the study period. The flowchart will also include the number of participants alive and available at each of the follow-up periods (6-month and 12-Month) as well as the number of participants lost to follow-up.

### 7.2 Baseline comparison

The baseline characteristics of villages, households and individuals are described below.

#### 7.2.1 Cluster characteristics

Description of the cluster characteristics will be presented by treatment group (Supplementary Table 1). Also, the characteristics of the BP management team members (family health instructors, village doctors and family leaders) in the intervention group will be presented in Supplementary Table 3. Categorical variables will be summarised by frequencies and percentages. Percentages will be calculated according to the number of clusters with available data. Continuous variables will be summarised by using mean and standard deviation (SD), or median and interquartile range (Q1-Q3).

#### 7.2.2 Household characteristics

The number of residents and the percentage of low-sodium salt usage are collected at the household level and will be presented by treatment group (Supplementary Table 1). The number of residents is continuous and will be summarised by mean and standard deviation (SD), or median and interquartile range (Q1-Q3). The percentage of low-sodium salt usage is categorical variable and will be summarised by frequencies and percentages.

#### 7.2.3 Individual characteristics

Description of the individual characteristics will be presented by treatment groups (Table 1). Additionally, the awareness, treatment and control rates of hypertension in participants with hypertension (defined as either a prior diagnosis of hypertension or a baseline blood pressure ≥140/90 mmHg) will be shown in Supplementary Table 2. Categorical variables will be summarised by frequencies and percentages. Percentages will be calculated according to the number of participants for whom data are available. Continuous variables will be summarised using mean and standard deviation if approximately normally distributed, or median and interquartile range (Q1-Q3) if normality is violated. No statistical test will be performed on baseline characteristics. No adjustment for clustering will be applied when summarising baseline characteristics. Baseline measurements for all participants are recorded in the baseline registration form and will be tabulated for the variables listed below:

- Anthropometrics: age (years), sex and BMI (kg/m^2^)
- Demographics: education
- Blood pressure: systolic blood pressure (mmHg) and diastolic blood pressure (mmHg)
- Medical history: hypertension, diabetes, dyslipidemia, coronary artery disease, stroke, atrial fibrillation, heart failure, chronic kidney disease, chronic obstructive pulmonary disease, asthma and malignant tumor
- Lifestyle factors: smoking, drinking, physical activity status (GPAQ scale) and dietary habits (Mini-EAT scale)

**Table 1.**
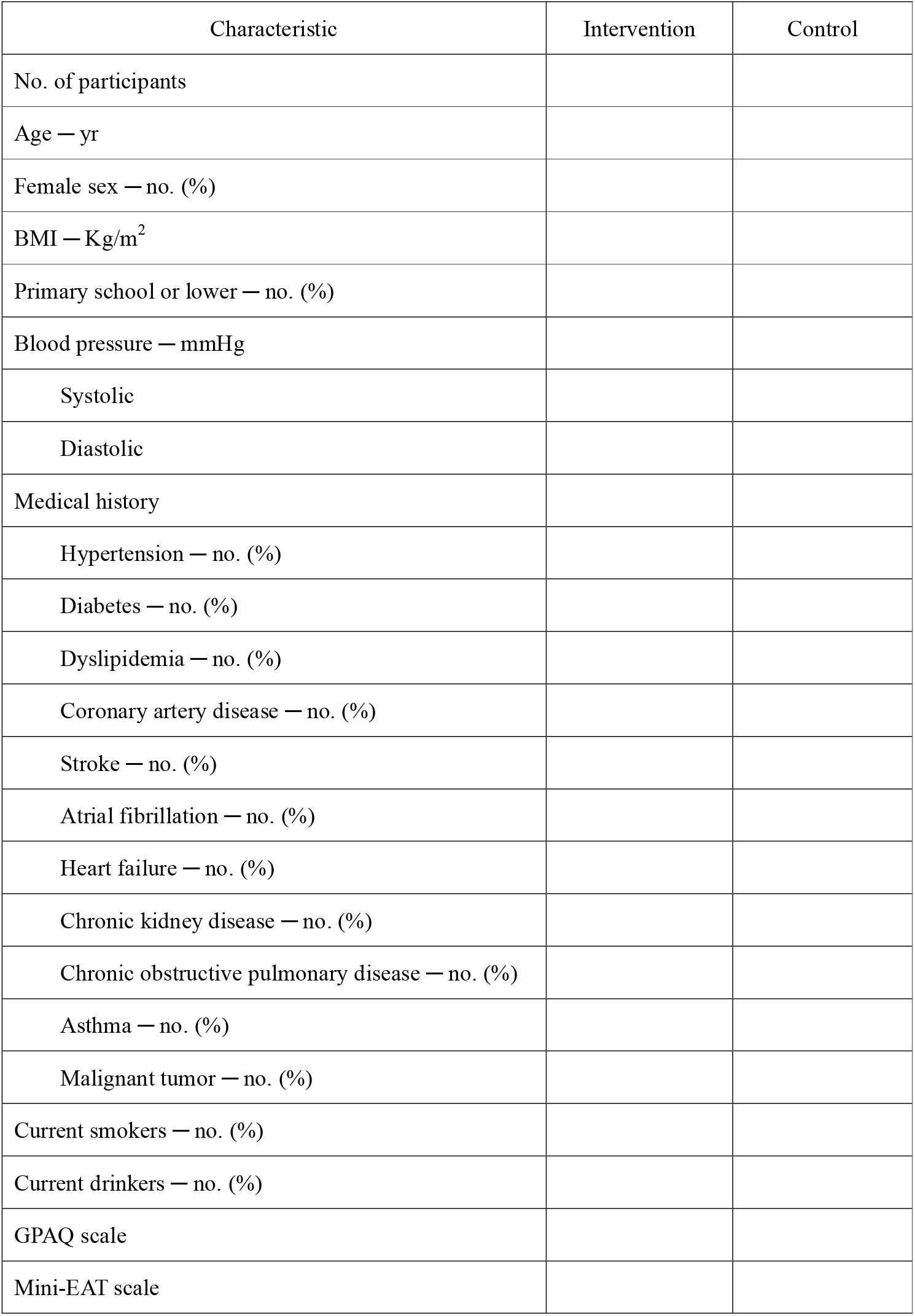
Baseline characteristics of trial participants.

| Characteristic | Intervention | Control |
| --- | --- | --- |
| No. of participants |  |  |
| Age — yr |  |  |
| Female sex — no. (%) |  |  |
| BMI — Kg/m<sup>2</sup> |  |  |
| Primary school or lower — no. (%) |  |  |
| Blood pressure — mmHg |  |  |
| Systolic |  |  |
| Diastolic |  |  |
| Medical history |  |  |
| Hypertension — no. (%) |  |  |
| Diabetes — no. (%) |  |  |
| Dyslipidemia — no. (%) |  |  |
| Coronary artery disease — no. (%) |  |  |
| Stroke — no. (%) |  |  |
| Atrial fibrillation — no. (%) |  |  |
| Heart failure — no. (%) |  |  |
| Chronic kidney disease — no. (%) |  |  |
| Chronic obstructive pulmonary disease — no. (%) |  |  |
| Asthma — no. (%) |  |  |
| Malignant tumor — no. (%) |  |  |
| Current smokers — no. (%) |  |  |
| Current drinkers — no. (%) |  |  |
| GPAQ scale |  |  |
| Mini-EAT scale |  |  |

### 7.3 Follow-up assessments

All assessments performed and interventions received during follow-up will be described by treatment group. The assessment will be performed at 6-month and 12-month home visits. The distribution of SBP at baseline, 6-month and 12-month will be shown in Supplementary Figure 1. The characteristics of antihypertensive medication usage, Global Physical Activity Questionnaire (GPAQ) scale, Mini-Eating Assessment Tool (Mini-EAT) scale[6], weight and number of antihypertensive medications will be shown in Supplementary Table 4-6. The distribution of self-reported SBP levels via the WeChat mini-program “Ruyang Heart Health” is presented in Supplementary Table 8, which reports data by month throughout the trial period.

During the 6-month visits, in addition to collecting village and individual information, any CVD events and adverse events will be recorded. The acceptable tolerance in the home visits is -7 days to +30 days. All assessments will be performed by independent assessors (masked to randomization status). More details of the follow-up assessments are available in the protocol. No formal statistical tests are planned for these variables.

### 7.4 Adherence and protocol deviations

Multifaceted intervention exposure is assessed based on implementation performance. We will evaluate the effectiveness of intervention by examining its main components of the multifaceted intervention (Supplementary Table 7). These variables are estimated along with corresponding 95% CI for village for the intervention arm based on the ITT population.

### 7.5 Analysis of the primary outcome

The primary outcome of this trial is the change in SBP from baseline to 6 months in all participants for comparison between the intervention group and control group (Table 2). The primary outcome will be analyzed by a mixed effect regression model, with change in SBP from baseline to 6 months included as the dependent variable, intervention allocation included as the independent variable of interest, township, age, sex, baseline SBP and history of hypertension as covariates to be adjusted, and village and family (nested within village) included as random effects with Gaussian distribution. All primary analysis models will include fixed effects for township, age, sex, baseline SBP, and history of hypertension. Other potential distributions within the exponential family will be considered for the primary outcome in case of non-normality. Figure 2 show the distribution of SBP at baseline and 6 months in both the intervention and control groups.

**Table 2.**
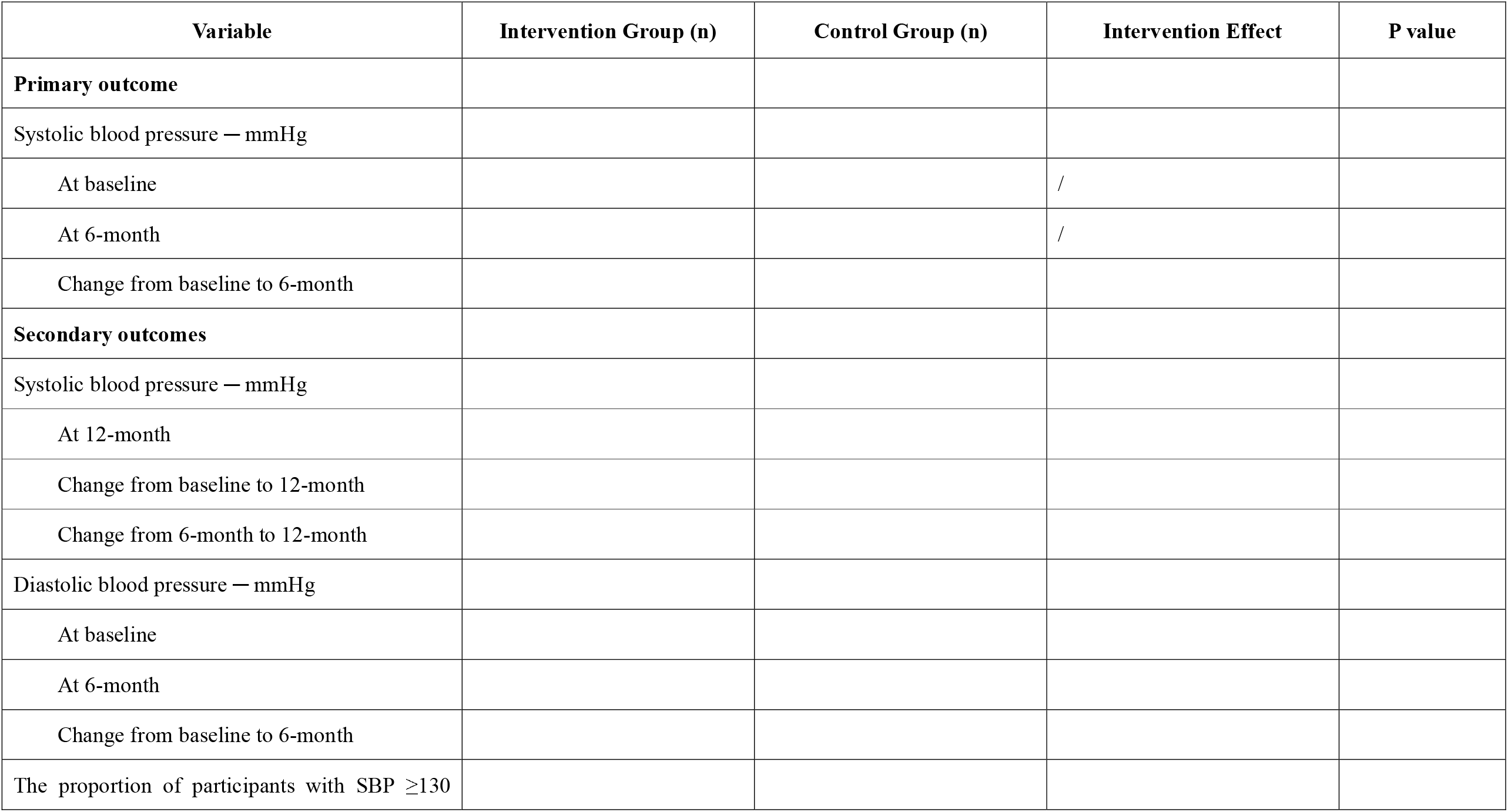
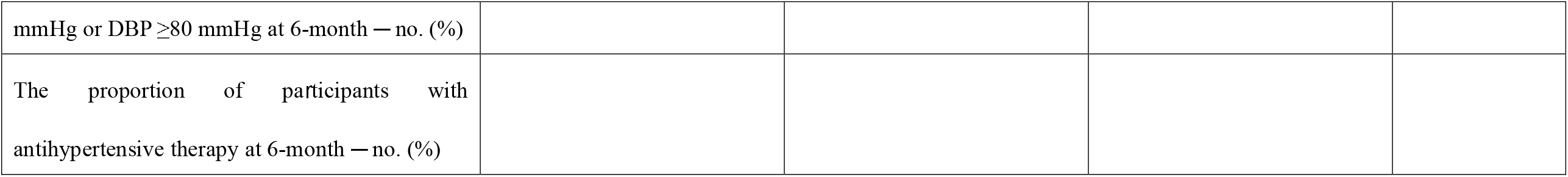
Intervention effect on primary and secondary outcomes.

**Figure 2.**
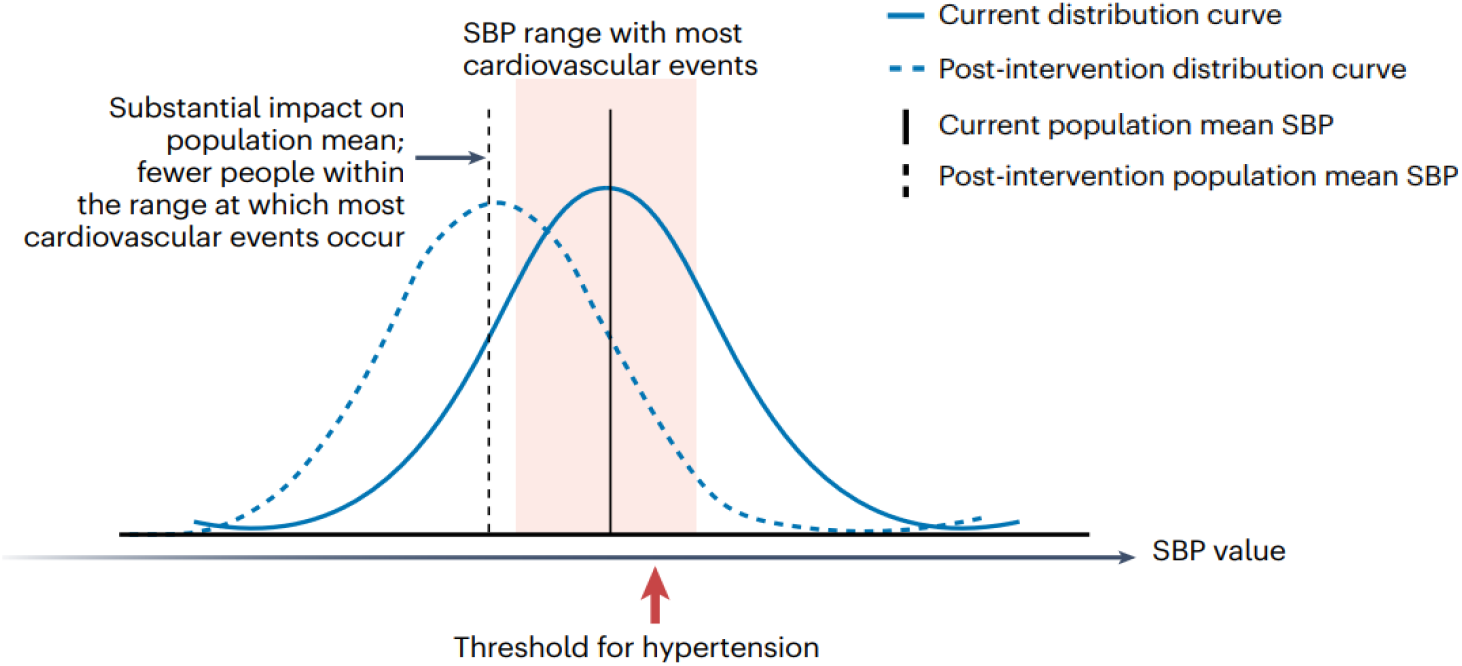
The distribution of systolic blood pressure from baseline to 6-month in the intervention and control group.

#### Main analysis (Individual-level)

Individual-level analysis will be performed to compare the change of SBP from baseline to 6 months in all participants between the intervention group and control group (as described in detail in section 5.1). The main analysis will be performed in the ITT population. Treatment allocation (multifaceted interventions vs control group) will be included as groups to be compared. The effect of the intervention will be presented as the difference of SBP change (6 months vs baseline) between the intervention and control group, and its 95% CI.

In the mixed effect regression model below,

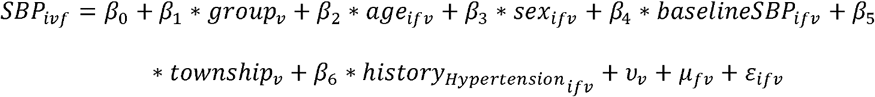

*i* refers to the participant, *v* refers to the village, and *f* refers to the family. *SBP*_*ifv*_ is the SBP change at 6 months relative to the baseline for the ith participant in the fth family of the vth village, *group*_*v*_ is the treatment allocation for the vth village (*o* =1 if the ith village is randomized to the intervention group, and *group*_*v*_ =0 if it is randomized into the control group), *age*_*ifv*_,*sex*_*ifv*_,*baselineSBP*_*ifv*_,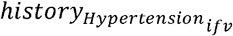 stand for the age, sex, baseline SBP and history of hypertension for the ith participants in the fth family of the vth village, respectively, and *township*_*v*_ refer to the town where the vth village located at. *υv* is the village random effect, where we assume that it follows a normal distribution with mean 0 and fixed variance,

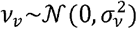

and its inclusion will take into account the clustering effect within village due to the cluster randomization. *µ*_*fv*_ is the nested random effect of family, where we assume it follows a normal distribution with mean 0 and fixed variance,

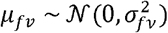

and its inclusion will take into account the clustering of participants within a family nested within a village. Since the number of participants within a family is relatively small, the estimation of 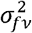 could be numerically unstable. When 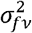 could not be estimated, a linear mixed effect model with the same set of fixed effects and village random effect will be conducted.

The treatment effect is estimated by 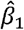, which represents the average difference of SBP change in the treatment group relative to the control group, adjusting for township, age, sex, baseline SBP, history of hypertension and village clustering effect. The 95% CI of the treatment effect captures the 95% CI of 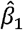, and can be calculated by 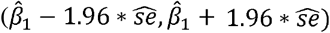, where 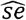is the estimated standard error of 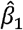. The statistical significance of the treatment effect is captured by the p value and 95% CI of 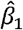.

Results will be presented by the estimated difference of SBP change between the treatment and control group, with estimated 95% CI. If the estimated difference in SBP change exceeds 0 and the 95% CI does not include 0, then we conclude that the multifaceted intervention decreases SBP compared to the control group with statistical significance.

### 7.6 Subgroup analyses

Eight subgroup analyses will be carried out, irrespective of whether there is a significant treatment effect on the primary outcome. These subgroup analyses will be performed in the ITT population.

Subgroups based on participant characteristics are defined as follows:

- Age (< 65 vs. ≥ 65 years);
- Sex (male vs. female);
- Education (≤ Primary school vs.>Primary school);
- History of hypertension (yes vs. no);
- Baseline SBP (< 140 mmHg vs. ≥ 140 mmHg);
- Baseline DBP (< 90 mmHg vs. ≥ 90 mmHg);
- Baseline Mini-EAT scale (< median vs. ≥ median);
- History of atherosclerotic cardiovascular disease (ASCVD) status (yes vs. no);
- Family size (2 vs. ≥3 family members within a family)
- Township socio-economic level (industrial enterprise count <40 vs. ≥40)

The analysis for each subgroup will be performed by first stratifying the ITT population according to each subgroup variable, and then calculating the difference of SBP change (and 95% CI) within each stratum. If the number of patients within a subgroup is too small, e.g. less than 5% of the overall population, the analyses may not be performed or the subgroup levels may be combined. The results will be displayed on a forest plot, including the point estimate and 95% confidence interval associated with the difference of SBP change in each stratum of the subgroup (Figure 3).

**Figure 3.**
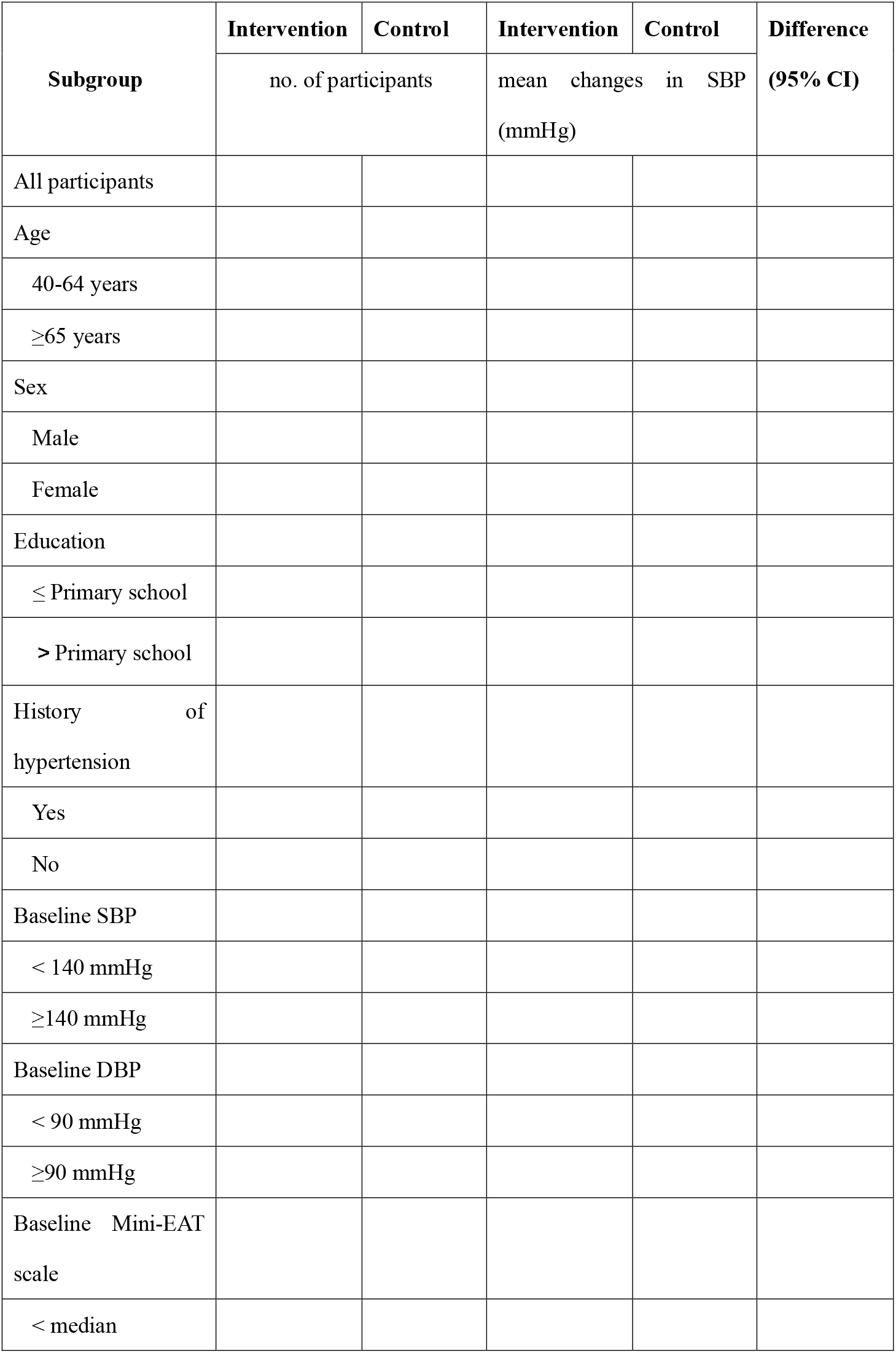
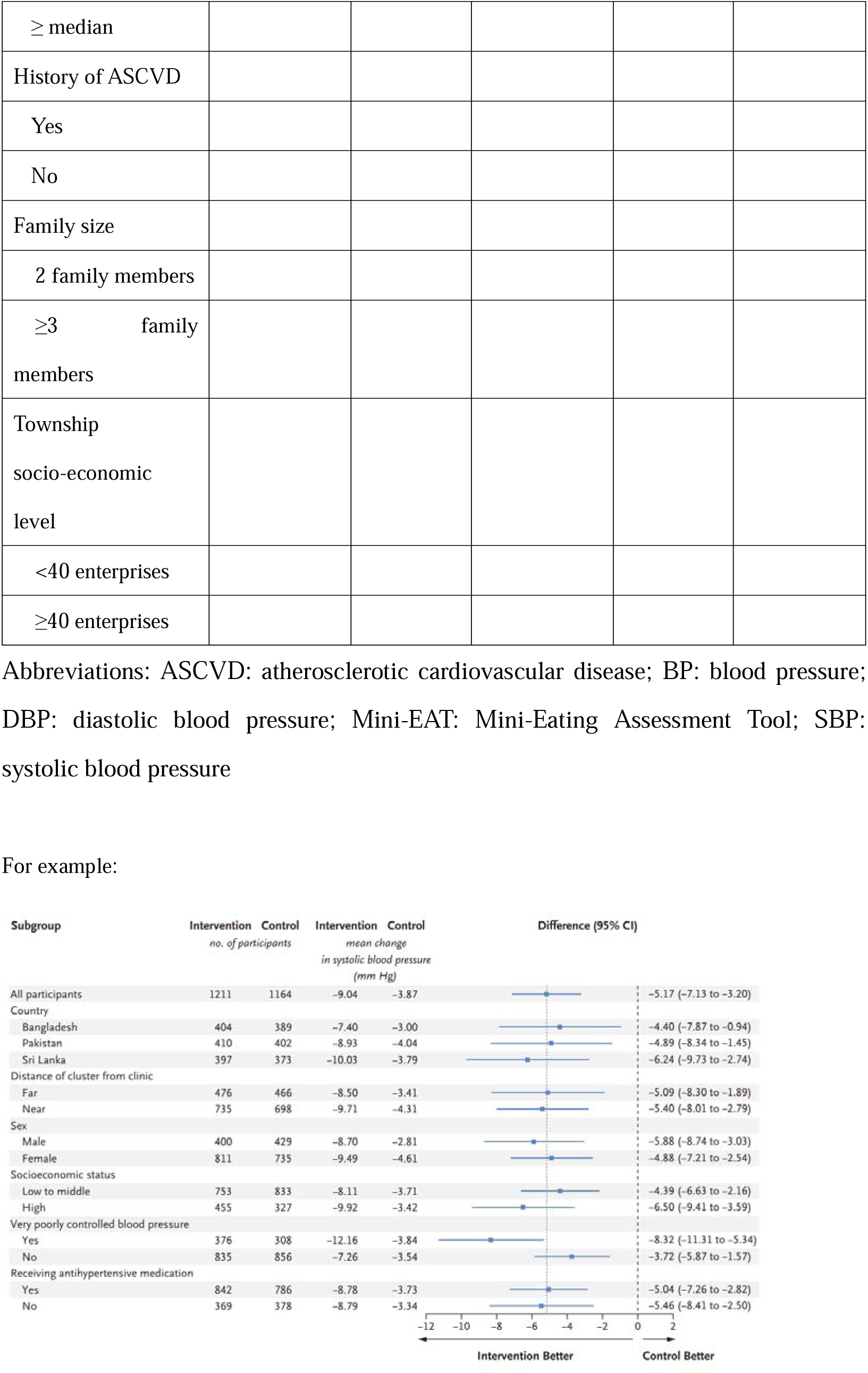
Subgroup analyses for change in systolic blood pressure at 6 months from baseline based on participant characteristics at baseline.

### 7.7 Adjusted analysis

Adjusted analyses of the primary outcome will be performed using individual-level data by adding the following covariates to the main analysis model (see section 7.5): BMI, education, history of diabetes, dyslipidemia, atrial fibrillation, heart failure and chronic kidney disease. The adjusted treatment effect will be reported as the difference of SBP change and 95% CI (Supplementary Table 9).

### 7.8 Sensitivity analyses

We will also perform a village-level analysis based on village-level mean change in systolic blood pressure using a linear model with fixed effects for township, age, sex, baseline SBP and history of hypertension; and a family-level analysis based on family-level mean change in SBP using a linear mixed effect model with fixed effects of township, age, sex, baseline SBP and history of hypertension, and random effect of village (Supplementary Table 10).

### 7.9 Analysis of missing data

The primary analysis is planned with no imputation for missing data.

The primary analysis, based on a likelihood based mixed effect regression model, is valid under the missing at random (MAR) assumption [7]. The MAR assumption indicates that whether the outcome of a participant is missing is independent of the unobserved outcome values after accounting for the appropriate observed outcome and covariates in the model. In order to evaluate the robustness of the findings to the MAR assumption, sensitivity analyses will be performed under varying assumptions for data considered likely to be missing under MAR, as well as missing not at random (MNAR). MNAR indicates that whether the outcome of a participant is missing depends on the unobserved values and cannot be predicted solely based on the participant’s observed data. Several types of statistical models have been proposed to analyze clinical study data under such assumptions.

The approach that will be implemented for this study is the use of multiple imputations for the primary outcome. The imputed outcome will be modelled using mixed effect regression model in the same way as the primary analysis.

It is expected that the majority of missing data will be caused by participants data collected outside the acceptable assessment window (−7 days to +30 days) lost to follow-up, which will result in a monotone missing pattern (once data from a participant is missing, his/her data at the next follow-up would also be missing). It is also expected that a small number of participants will be missing at the first follow-up, but non-missing at the second follow-up, where data at the first follow-up will be imputed before imputing the monotone missing data.

### 7.10 Per-protocol analysis

No per-protocol analysis will be performed.

### 7.11 Analysis of secondary outcomes (Individual-level analysis)

All secondary outcome analyses described in this section will be performed in the ITT population (Table 2). Continuous secondary outcomes will be analyzed using a similar mixed effect regression model with the primary outcome. Appropriate distributions within the exponential family will be considered when these outcomes have skewed distributions. Categorical secondary outcome will be analyzed using a generalized linear mixed effect regression model with logit link function, where binary secondary outcome is included as the dependent variable, intervention allocation included as the independent variable of interest, age, sex, baseline SBP, township and history of hypertension as covariates to be adjusted, and village and family included as the random effects with Gaussian distribution. In case of the family random effects not being estimated, (generalized) linear mixed effect regression model with the same set of fixed effects and village random effect will be conducted.

### 7.12 Analysis of adverse events and cardiovascular events

Adverse events, including SAEs and AESIs, will be summarized by the number of occurrences. This will be done overall and by event category according to Medical Dictionary for Regulatory Activities (MeDRA) system organ classes and preferred terms. Cardiovascular events, including cardiovascular mortality, heart failure hospitalization, non-fatal myocardial infraction and non-fatal stroke, will also be summarised. Both adverse and cardiovascular events will be summarised by intervention arm (Supplementary Table 11), with no formal statistical tests applied. The safety analysis will be performed on the ITT population.

### 7.13 Analysis of program delivery cost

Intervention delivery costs are based on the activity-based costing exercise conducted prospectively during the trial. The costs of implementing the multifaceted intervention will be assessed at the 6 months (Supplementary Table 12). Additionally, we will estimate the incremental costs of scaling up Healthy Family Program to all eligible adults aged 40 to 80 in rural areas in China from the perspective of the program implementer in a separated economic analysis.

## Data Availability

All data produced in the present study are available upon reasonable request to the authors

## 9 Appendix 1: Proposed tables and figures

## Supplementary materials

**Supplementary Table 1.** Baseline characteristics of trial villages and households.

| Characteristic | Intervention | Control |
| --- | --- | --- |
| **Villages** | N= | N= |
| Total No. of households |  |  |
| Total No. of residents |  |  |
| Annual income per resident |  |  |
| <30,000 yuan |  |  |
| (30,000-50,000] yuan |  |  |
| (50,000-100,000] yuan |  |  |
| >100,000 yuan |  |  |
| Distance from Ruyang County People's Hospital |  |  |
| **Households** | N= | N= |
| Mean number of family members per household |  |  |
| Use of low-sodium salt, n/N (%) |  |  |

**Supplementary Table 2.** Awareness, treatment, and control rates of hypertension at baseline.

| Characteristic | Intervention | Control |
| --- | --- | --- |
| Hypertension* — no. (%) |  |  |
| Awareness, n/N (%) |  |  |
| Use of antihypertensive treatment, n/N (%) |  |  |
| Controlled (SBP<130 and DBP<80 mmHg), n/N (%) |  |  |

\*Hypertension is defined as either a prior diagnosis of hypertension or a baseline blood pressure $\geq 140/90$ mmHg

**Supplementary table 3.**
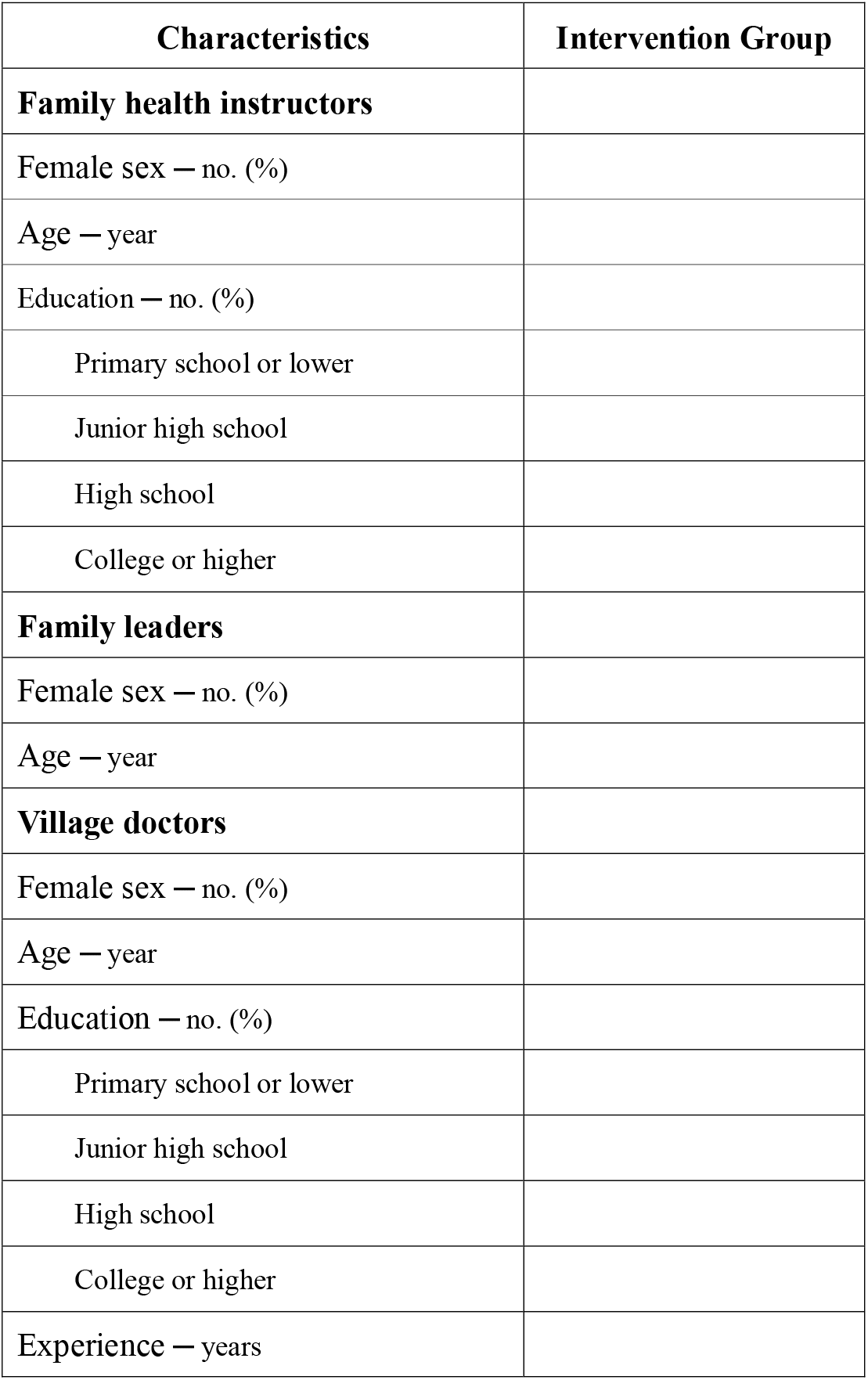
Characteristics of blood pressure management team members in the intervention group.

**Supplementary Table 4.**
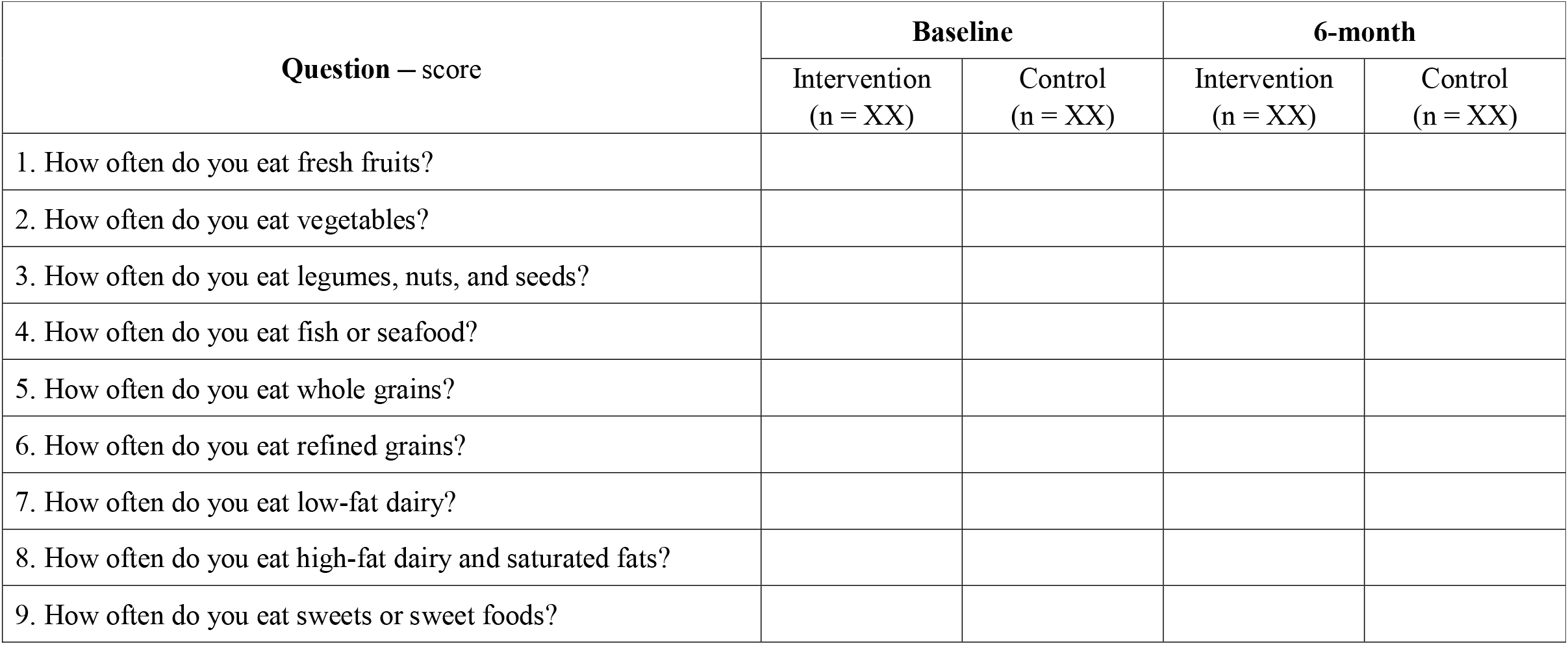
The Mini-Eating Assessment Tool (Mini-EAT) scale in both groups.

**Supplementary Table 5.**
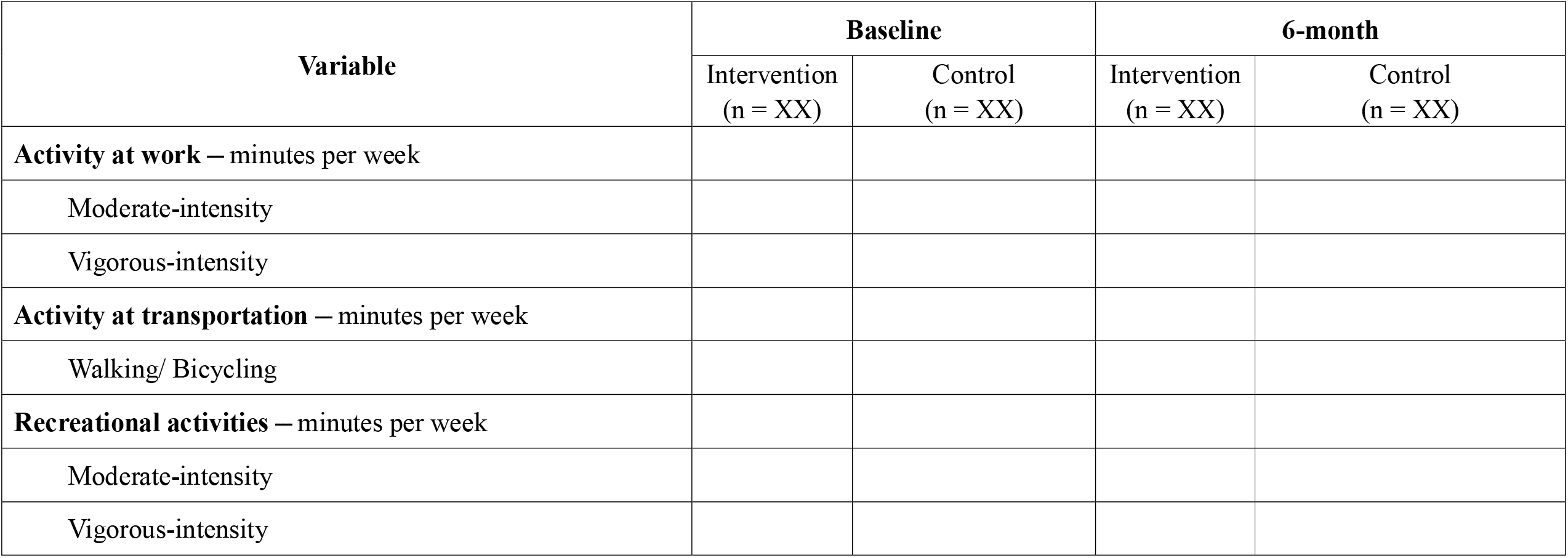
The Global Physical Activity Questionnaire (GPAQ) scale in both groups.

**Supplementary Table 6.**
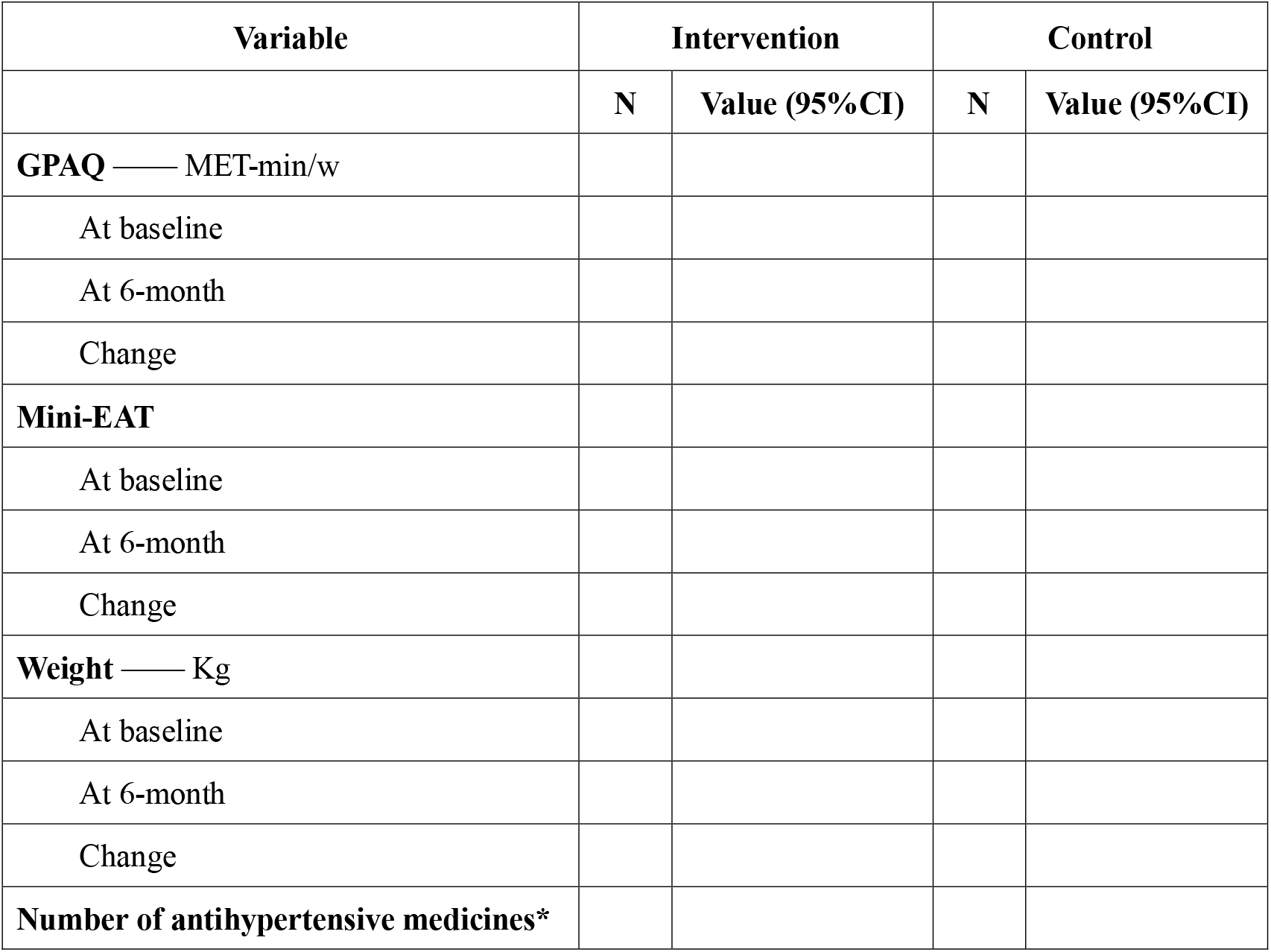
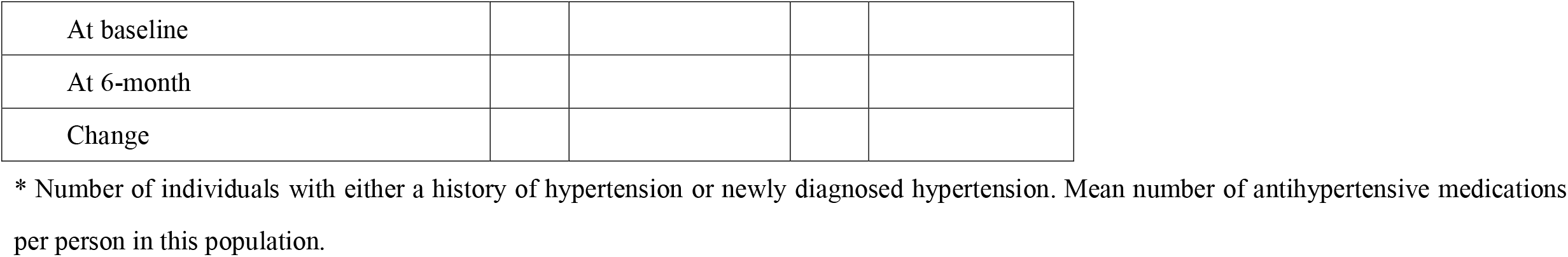
Summary of antihypertensive medication usage, weight, Global Physical Activity Questionnaire (GPAQ) scale and Mini-Eating Assessment Tool (Mini-EAT) scale.

**Supplementary Table 7.** Information of implementation in the intervention group.

| Variable | The first month | The fourth month |
| --- | --- | --- |
| Education for Health Family Instructors by research team | / | / |
| Attendance Rate of Health Family Instructors (%) |  |  |
| Session Duration — minutes |  |  |
| Education for participants by Health Family Instructors | / | / |
| Attendance Rate of Family Leaders (%) |  |  |
| Session Duration — minutes |  |  |
| Healthy lifestyle pamphlets distribution (%) |  |  |
| Sodium substitute salt distribution (%) |  |  |
| Weighing scale distribution (%) |  | / |
| Physical exercise organization (times/week)* |  |  |
| Blood pressure monitor distribution (%) |  | / |
| Training on the use of Blood Pressure Management Platform (%) |  | / |
| Education of Blood Pressure Management for village doctors (%) |  | / |

\*Average weekly frequency of organized physical exercises per village during the intervention period

**Supplementary Table 8.**
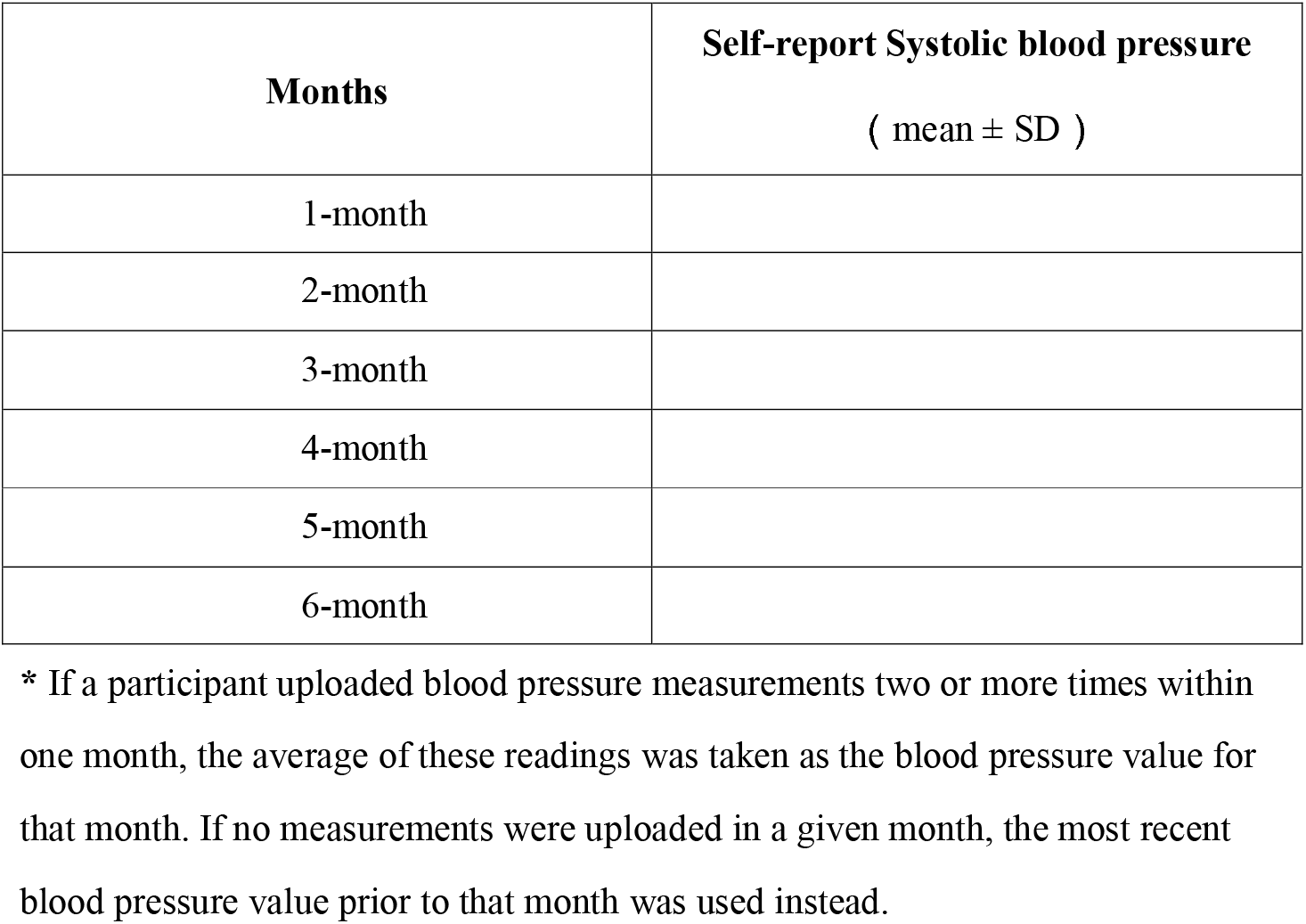
The distribution of different levels of self-report systolic blood pressure in the intervention group during the trial period*.

**Supplementary Table 9.**
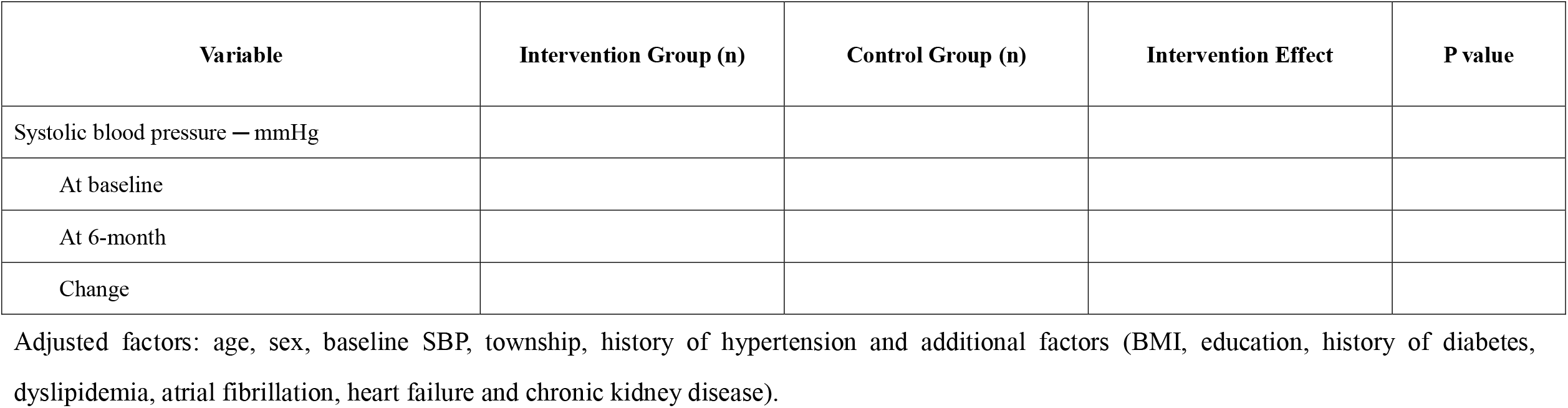
Intervention effect on primary outcome after additional adjustment (sensitivity analysis)

**Supplementary Table 10.**
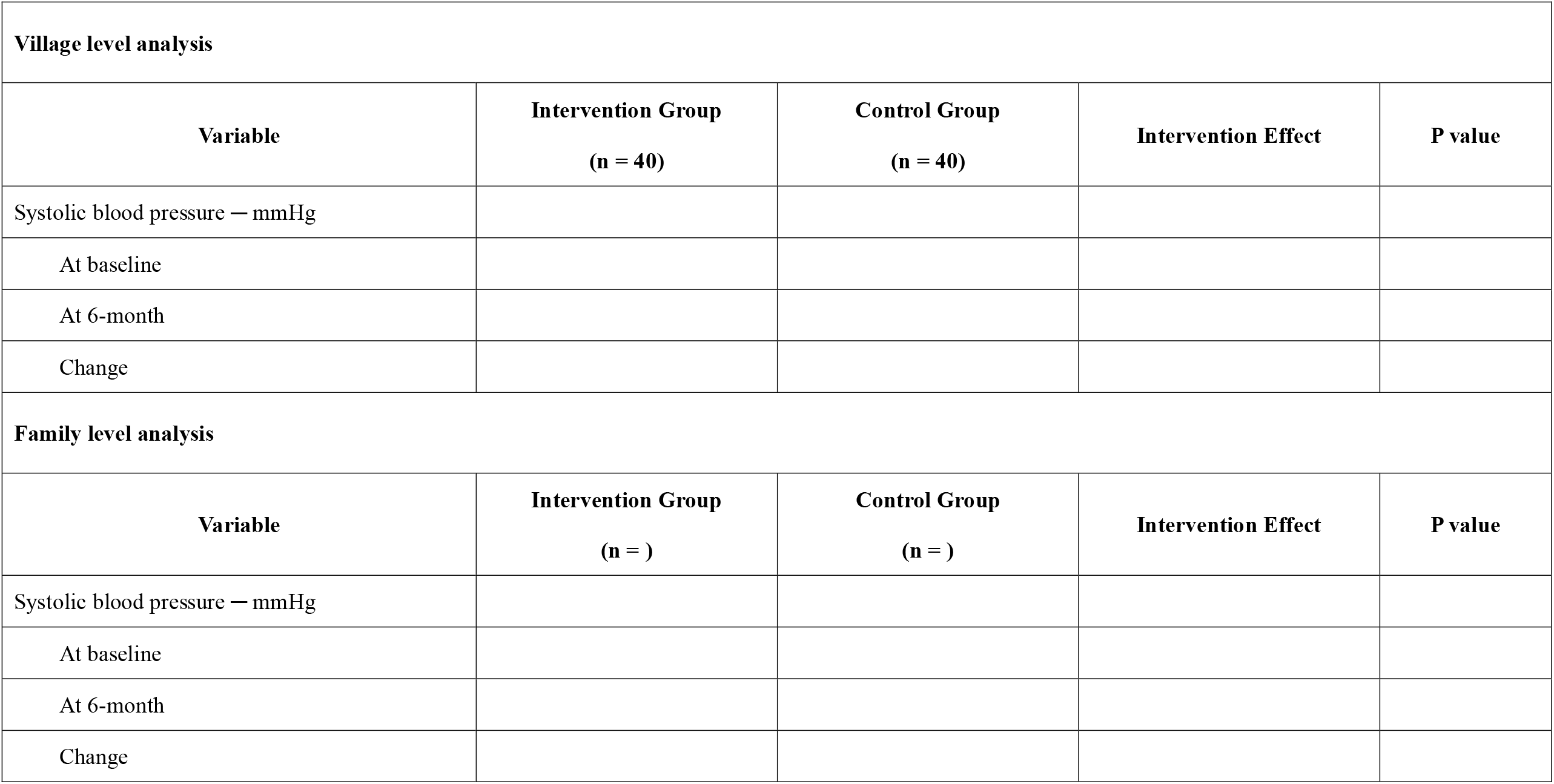
Cluster-level analysis for intervention effect on blood pressure (sensitivity analysis)

**Supplementary Table 11.** Adverse events and cardiovascular events at 6 months.

| Variable | At 6 months |  |
| --- | --- | --- |
|  | Intervention Group (n) | Control Group (n) |
| **Serious Adverse events (SAEs)** |  |  |
| **Adverse Events of Special Interest** |  |  |
| Hypotension |  |  |
| Syncope |  |  |
| Hyponatremia |  |  |
| Hyperkalemia |  |  |
| Injurious falls |  |  |
| Acute kidney injury |  |  |
| **Cardiovascular events — no. (%)** |  |  |
| Cardiovascular mortality |  |  |
| Heart failure hospitalization |  |  |
| Non-fatal myocardial infraction |  |  |
| Non-fatal stroke |  |  |
| **All-cause mortality** |  |  |

**Supplementary Table 12.** The key cost components of implementing the multifaceted intervention in this program.

| Activities | Costs (CNY) |
| --- | --- |
| **Administration** |  |
| Cost of transportation, accommodation and meals for researchers visiting to study sites |  |
| Subsidies of transportation and meal for study coordinators visiting to study sites |  |
| Fees paid to study coordinators and management team in the intervention period |  |
| **Implementation of multifaceted interventions to participants** |  |
| Fees paid to family health instructors for performing their duties |  |
| Free provision of low-sodium substitutes |  |
| Free provision of weight monitors |  |
| Blood pressure monitoring application development and maintenance |  |
| Free provision of blood pressure monitors |  |
| Training material: printing of posters, manuals, and handbooks |  |
| **Total Cost (CNY)** |  |
| Cost per person (4001 participants) |  |

**Supplementary Figure 1.**
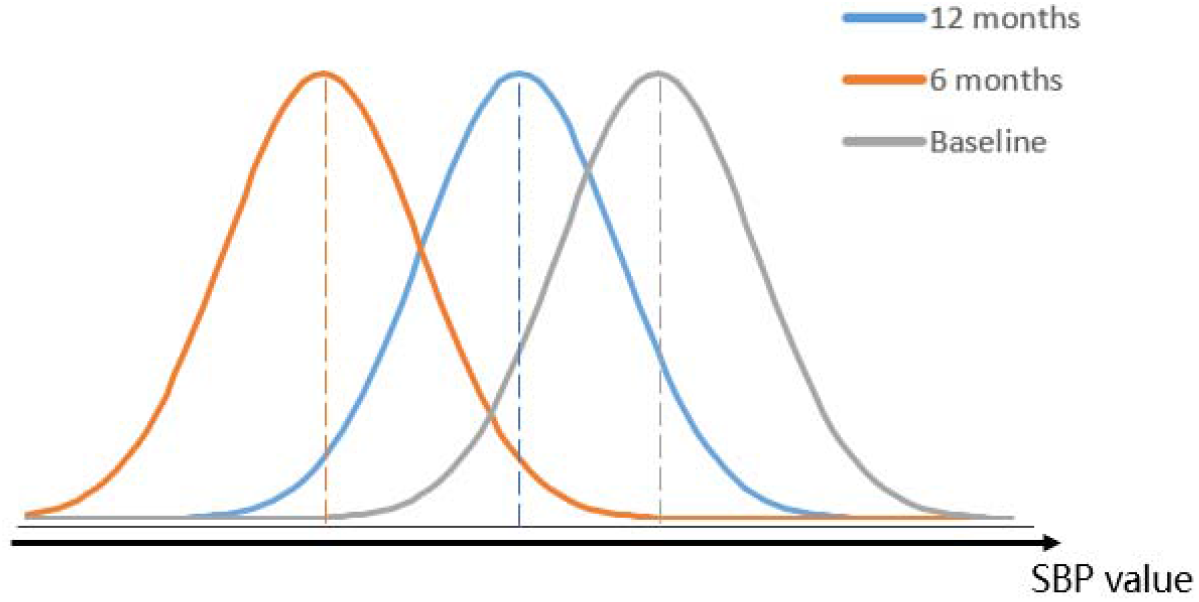
The distribution of systolic blood pressure at baseline, 6-month and 12-month in the intervention and control group.

